# Fractionated Potential-Guided Cryoablation Targeting Termination of Atrioventricular Nodal Reentrant Tachycardia While Avoiding the Compact Node Electrogram

**DOI:** 10.1101/2024.11.18.24317515

**Authors:** Shu Hirata, Koichi Nagashima, Ryuta Watanabe, Yuji Wakamatsu, Moyuru Hirata, Sayaka Kurokawa, Naoto Otsuka, Masanaru Sawada, Yasuo Okumura

## Abstract

**Background:** Fractionated potential (FP) ablation during atrioventricular nodal reentrant tachycardia (AVNRT), is an effective strategy that minimizes redundant radiofrequency (RF) applications. This study aimed to evaluate the utility of cryoablation targeting FPs to effectively terminate AVNRT while further minimizing redundant cryoapplications. Moreover, we observed what appeared to be compact AVN (cAVN) or proximal His potentials—tiny, dull potentials (TDPs) with continuity to the His potential during sinus rhythm (SR) and AVNRT—in the anteroseptal area. The second aim of this study was to explore the significance of those potentials.

**Methods:** Analyzed were 53 slow-fast AVNRT patients who underwent ablation procedures. Ultra-high resolution activation maps in the triangle of Koch were obtained during SR (n=34) and AVNRT (n=46). TDPs during SR and AVNRT in the anteroseptal area were identified and annotated using the LUMIPOINT Activation Search tool.

**Results:** FP areas were observed in 19 patients (56%) during SR and in 46 (100%) during AVNRT. This area corresponded to the AVNRT termination and/or successful ablation site in all, with peak numbers of 8.8±1.4 during AVNRT and 5.3±1.3 during SR. The number of ablation points was 3.6±1.5 for the FP-guided cryoablation (n=32) (Bonferroni corrected P<0.05 vs. anatomical RF; and P<0.05 vs. FP-guided RF), 5.4±2.1 for the FP-guided RF ablation (n=11) (P=0.0825 vs. anatomical RF), and 8.2±3.2 for the conventional RF ablation (n=10). Transient AV block occurred in 11 patients (21%). All AV block sites overlapped with the TDP area in the phase just before the His potential during AVNRT and SR, with a confidence setting of ≥24% (35[24-60]%). Conversely, in 42 patients without AV block, no ablation was performed in this area.

**Conclusion:** The FP-guided cryoablation strategy targeting AVNRT termination required fewer cryoapplications than RF ablation. The RF/cryo application in the TDP area during SR and AVNRT posed a risk of AV block.

## Introduction

Anatomical slow pathway (SP) ablation is an established treatment strategy for atrioventricular nodal reentrant tachycardia (AVNRT).^1^ Targeting the rightward inferior extension (RIE) of the SP offers a high success rate of 96% to 98%.^1,2^ Additionally, our previous research demonstrated that radiofrequency (RF) ablation targeting fractionated potentials, identified with ultrahigh-resolution mapping during AVNRT, is an effective strategy that minimizes redundant RF applications.^3^ Those results suggest that the anatomical and electrophysiological targets for SP ablation are well-defined. Despite this, atrioventricular (AV) block, necessitating a permanent pacemaker implantation, remains a significant complication, occurring in 0.2-2.4% of RF ablation cases.^4–6^ Cryoablation has been proposed as a promising alternative, particularly due to its potential to entirely reduce the risk of AV block.^7–9^ A key advantage of cryoablation is the reversibility of AV nodal (AVN) conduction if the cryoapplication is interrupted immediately after AV block occurs, allowing confirmation of AVNRT termination during the procedure.^9–11^ Given these considerations, the next step is to evaluate the utility of cryoablation targeting fractionated potentials to effectively terminate AVNRT while minimizing redundant cryoapplications in comparison to fractionated potential-guided RF applications during SR.

However, transient AV block still occurs during cryoablation in 25%, despite the absence of permanent AV block.^7,8^ This is because the electrogram reflecting the compact AVN (cAVN) activation has not been well defined. In this context, the ability to accurately visualize the cAVN is critical for safer ablation. Using the ultrahigh-resolution mapping technology, we have observed what appear to be cAVN or proximal His potentials—tiny, dull potentials with continuity to the His potential during sinus rhythm and AVNRT—in the anteroseptal area. However, those findings have not been fully clarified. Hypothesizing that those potentials represent the cAVN, the second aim of this study was to explore the significance of those potentials recorded by the ultrahigh-resolution mapping system.

## Methods

### Study Patients

Analyzed were 53 slow-fast AVNRT patients who underwent an ablation procedure in our institution between September 2017 and July 2024. The data collection and analysis were approved by the ethics committee of Nihon University Itabashi Hospital and that of each participating institution. Patients included had approved the use of their data for research purposes through an opt-out method.

### Electrophysiologic Study

Electrophysiologic studies were performed under conscious sedation: 4- to 10-pole electrode catheters were positioned in the high right atrium, right ventricular (RV) apex, and His bundle region through the right femoral vein and into the coronary sinus (CS) through the right jugular vein. Bipolar intracardiac potentials were digitally recorded (LabSystem PRO, Boston Scientific Corporation, Marlborough, MA) at a paper speed of 100–200 mm/s through a bandpass filter of 30–500 Hz. Bipolar pacing was performed at a current of 10 mA and pulse width of 2 ms. After exclusion of atrial tachycardia, by observation of a V-A-V response after RV overdrive pacing during the tachycardia or the presence of ventriculoatrial (VA) linking after differential atrial overdrive pacing,^12–14^ AVNRT was diagnosed if none of the following was present: (1) an advancement or delay of the His timing of >10 ms or termination of the tachycardia upon delivery of a scanned single premature ventricular contraction (PVC) during His-bundle refractoriness, (2) an uncorrected/corrected post-pacing interval (PPI) minus the tachycardia cycle length (TCL) of <115 ms/<110 ms, and (3) orthodromic His or septal ventricular capture during RV overdrive pacing at a cycle length of 10 ms to 30 ms less than the TCL.^13,15,16^ AVNRTs were further categorized as slow-fast AVNRT based on the observation of a His-atrial (HA) interval of ≤70 ms.^13,17^

### Mapping in the Triangle of Koch

An activation map focusing on the triangle of Koch was obtained with the use of a mini-basket catheter (64 mini-electrodes; 0.4 mm^2^, 2.5 mm center-to-center interelectrode spacing, IntellaMap Orion, Boston Scientific) and RHYTHMIA mapping system (Boston Scientific) during sinus rhythm (SR) and AVNRT (Figure 1A-B). The following criteria were used for the beating acceptance: (1) matching electrocardiographic morphology, (2) time stability of the electrograms, (3) respiratory stability, and (4) a maximal electrode to anatomical shell distance of 2 mm. Bipolar electrograms were filtered at 30 Hz to 300 Hz, and unipolar electrograms were filtered at 1 Hz to 300 Hz (default setting). The activation map was analyzed by means of the LUMIPOINT algorithm and complex activation module, which highlights regions showing both activation within the time-of-interest period and electrograms with multiple fractionated components. The highlighted areas are displayed based on the number of fractionated components, as adjusted by the peak slider. According to our previous publication, the number of peaks was initially set to 8.4 and then adjusted to ensure a highlighted area of approximately 1.0 cm² for each patient (Figure 1).^3^ A fractionated potential was considered absent if no area was highlighted even when the peaks were set to <5.0. If the atrial and ventricular potentials overlapped, the fractionation of the atrial potential was confirmed by separating the atrial and ventricular potentials using an atrial or RV extrastimulus.

**Figure 1.**
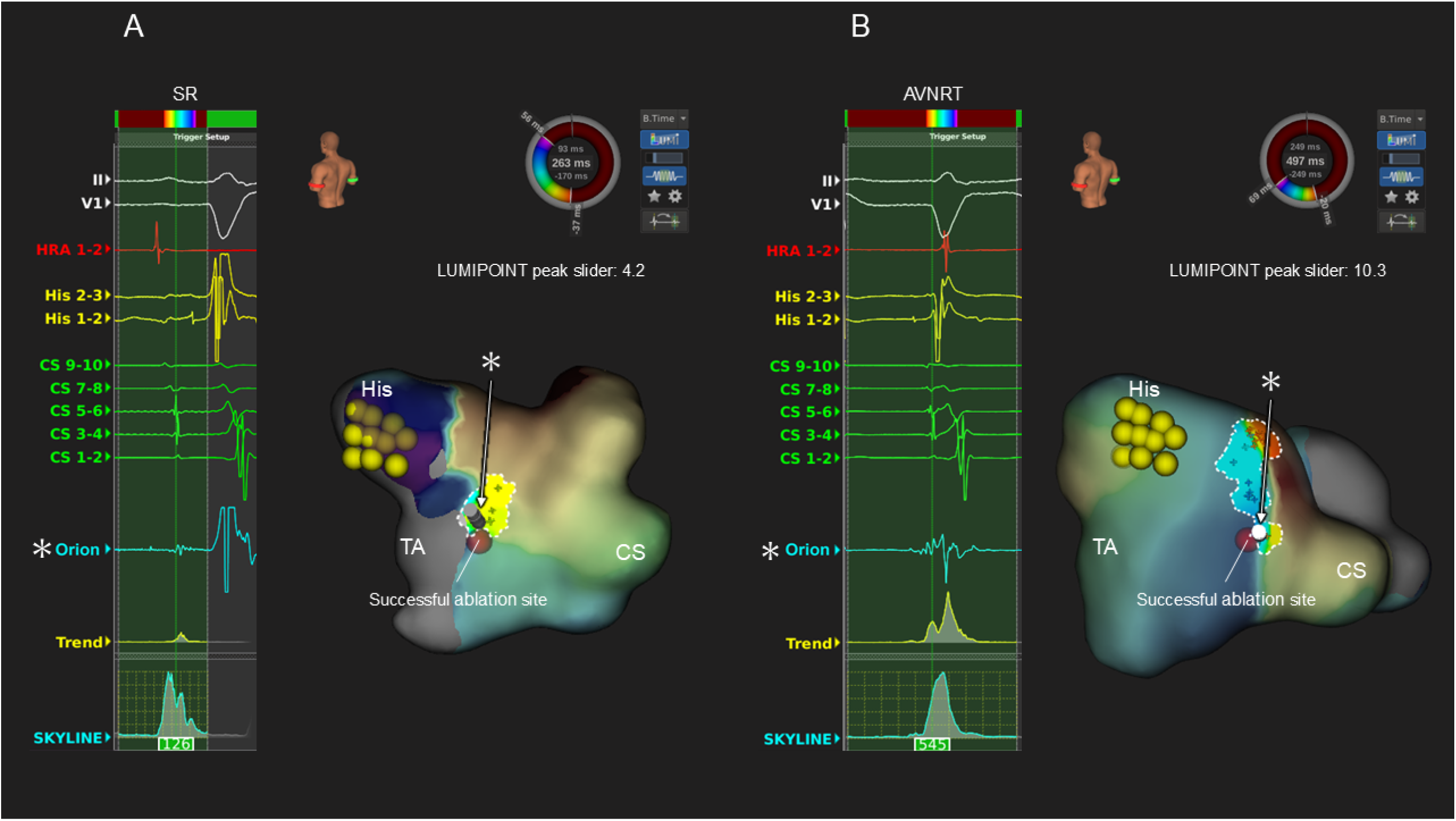
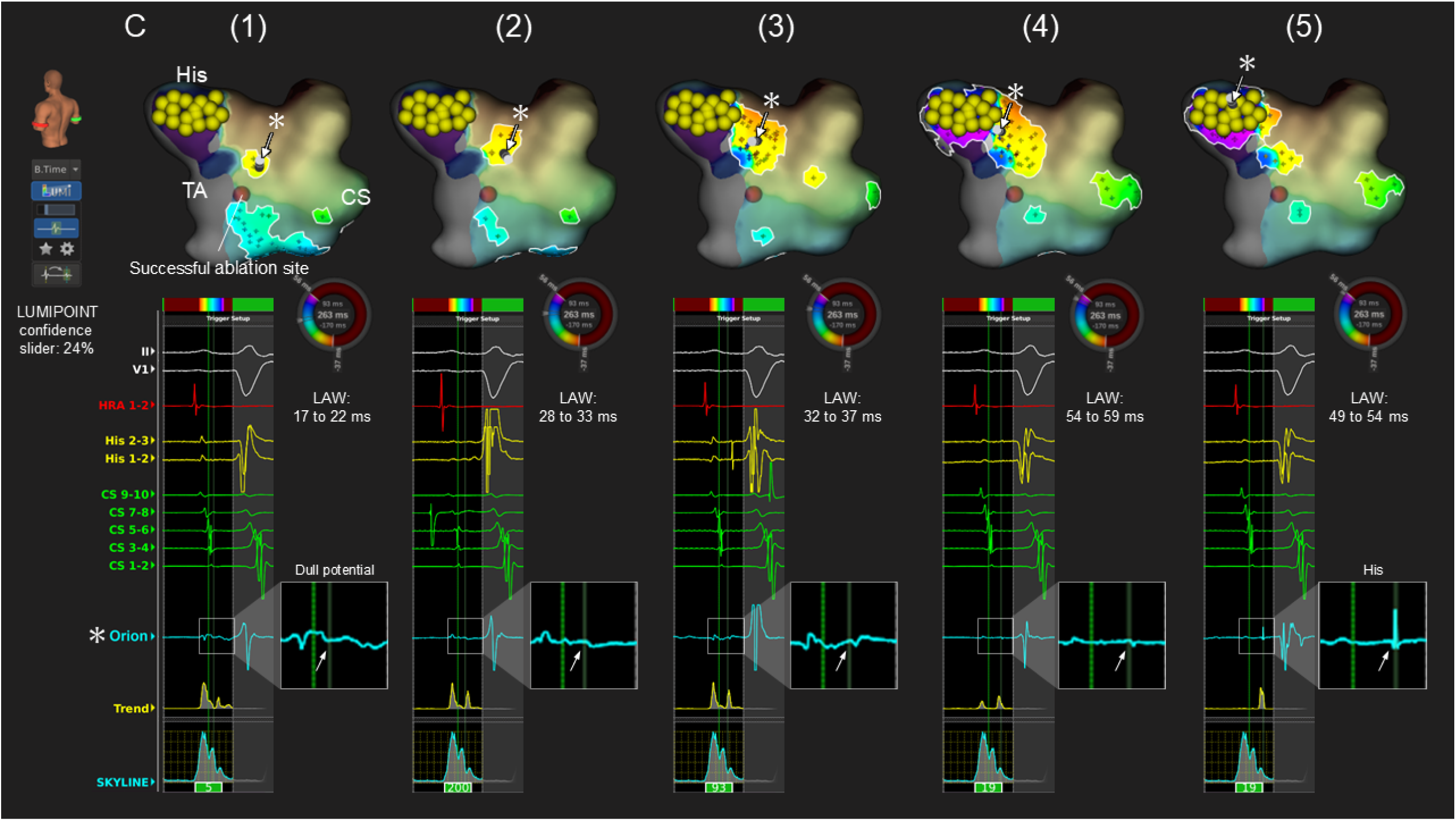
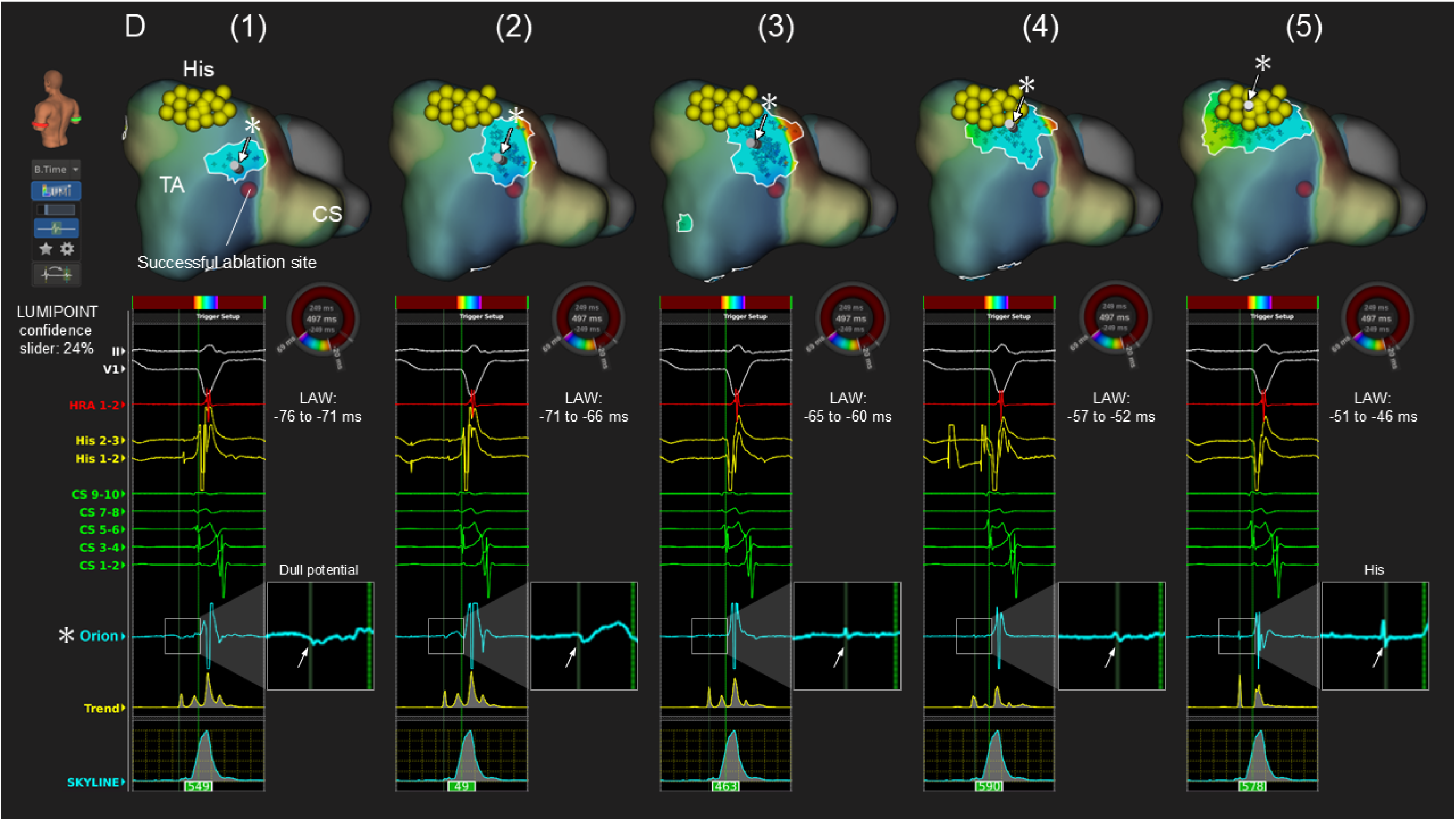
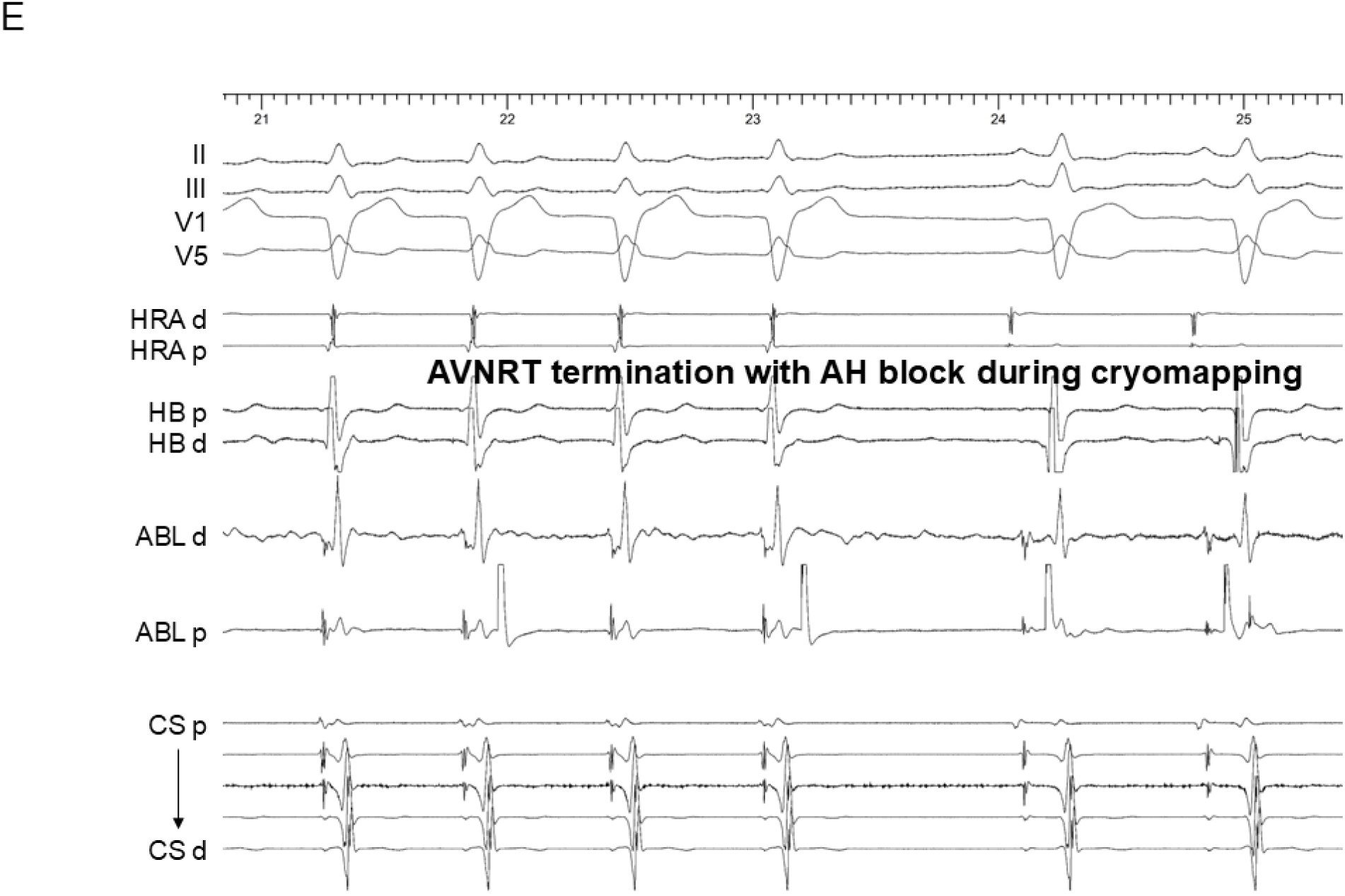
Representative case showing an activation map and local electrograms in the triangle of Koch. **(A)** An activation map and local electrograms during sinus rhythm (SR) and **(B)** atrioventricular nodal reentrant tachycardia (AVNRT). The fractionated potentials are highlighted during SR with peak numbers of 4.2 and during AVNRT with peak numbers of 10.3. The fractionated potential area corresponds to the successful AVNRT termination site. **(C)** The propagation of the annotated dull potentials during SR with confidence levels of 24% and **(D)** during AVNRT with confidence levels of 24%. By sliding the local activation window (LAW) by 5 ms, these potentials are observed propagating from the anteroseptal area to the His region. **(E)** Intracardiac electrograms of AVNRT termination during cryomapping. ABL = ablation catheter; CS = coronary sinus; d = distal; HB = His bundle; HRA = high right atrium; p = proximal; TA = tricuspid annulus.

Tiny, dull potentials with continuity to the His potential during SR and AVNRT in the anteroseptal area were identified and annotated using the LUMIPOINT Activation Search tool (Figure 1C-D). The activation of these potentials was visualized by reducing the confidence level, setting the LUMIPOINT Activation Window (LAW) to 5 ms, and sliding the LAW from the atrial activation timing in the anteroseptum to the His-bundle activation timing.^18^

### Ablation Procedure

A flow diagram of the ablation procedure details according to the three strategies including the fractionated potential-guided cryoablation, RF ablation, and conventional anatomical RF ablation, are shown in Figure 2. Cryoablation was performed in 32 patients. Cryomapping was applied to the fractionated signal area highlighted by the LUMIPOINT module using a 7Fr 6-mm-tip cryoablation catheter (Freezor Xtra, Medtronic, Minneapolis, MN) during AVNRT or during SR if AVNRT could not be reproducibly induced, with a cryocatheter tip temperature of −30°C for a maximum duration of up to 60 seconds. If AVNRT termination or the elimination of a dual AVN physiology was observed during cryomapping, cryoablation was then performed at the same site at a catheter tip temperature of −80°C for 240 seconds. If cryomapping failed to terminate the AVNRT, it was then attempted at another site. After the targeted lesion thawed to body temperature, an additional 240-second cryoapplication was performed at the same site. If AV block occurred, the cryoapplication was immediately interrupted. Following each cryoapplication, the dual AV nodal physiology and AVNRT inducibility were reassessed, with or without an isoproterenol infusion. If AVNRT remained inducible, the cryoapplication was gradually extended superiorly in the area highlighted by the LUMIPOINT module. Acute success was defined as the complete elimination of the slow pathway (SP) or the presence of an AH jump with or without a single AVN echo, in addition to the noninducibility of AVNRT. The site of successful ablation was defined as the location where the cryoapplication terminated the AVNRT or rendered it non-inducible.

**Figure 2.**
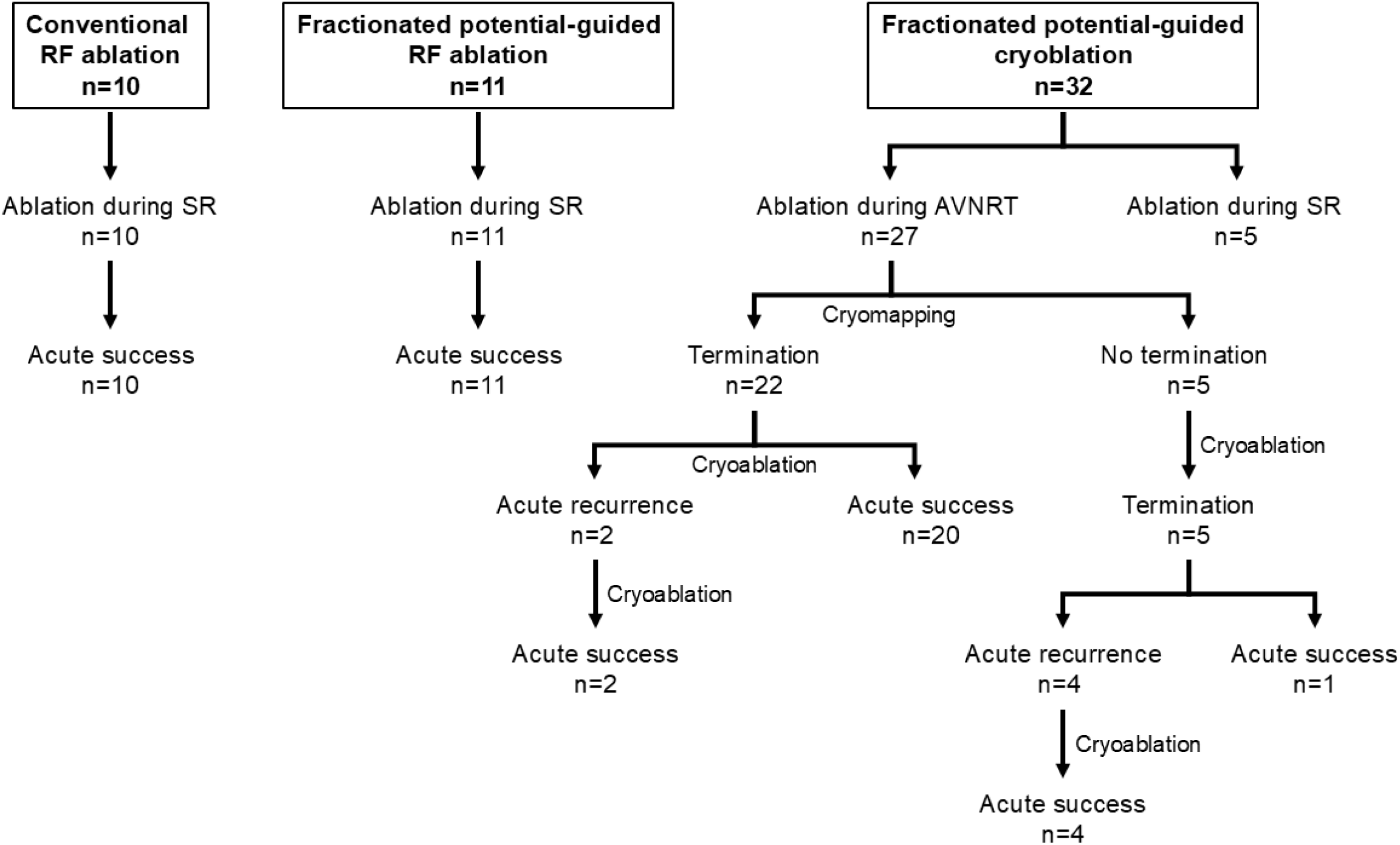
Flow diagram of the ablation procedure details according to the three strategies. AVNRT = atrioventricular nodal reentrant tachycardia, RF = radiofrequency, SR = sinus rhythm.

### Radiofrequency Ablation

In 10 of the 21 patients who received RF ablation, RF was applied by the conventional approach,^19,20^ where an atrial/ventricular electrogram ratio of <0.5 was obtained during sinus rhythm with a 4- or 4.5-mm open-irrigated catheter (INTELLANAV OI or INTELLANAV OI MiFi or INTELLANAV STABLEPOINT; Boston Scientific) with an output of 25 W to 35 W and used in the temperature-control mode at a target temperature of 55°C and a Maestro™ 4000 RF generator (Boston Scientific) used in conjunction with a MetriQTM irrigation pump (Boston Scientific). In the remaining 11 patients, RF energy was initially applied only to the fractionated potential area highlighted by the LUMIPOINT module.^3^ The RF duration was set for up to 60 seconds until junctional rhythm was elicited. If transient AV block occurred, the RF delivery was immediately stopped. The RF application was gradually extended to the higher area unless any junctional rhythm was seen during the energy delivery or AVNRT was still inducible. If a junctional rhythm was frequently seen, the dual AVN physiology and AVNRT inducibility were reassessed with/without an isoproterenol infusion. Acute success was defined as a complete SP elimination or AH jump-up with/without a single AVN echo, in addition to the noninducibility of AVNRT. Unless acute success was achieved, the RF energy was delivered in an area higher than the previous ablation area. A site of successful ablation was defined as the last site of the RF application where the AVNRT was rendered non-inducible.

### Statistical Analysis

Continuous variables are presented as the mean ± SD or as medians with interquartile ranges in parentheses, depending on the data distribution. To compare the continuous variables, either a Student’s t-test or Mann-Whitney U test was applied, based on the distribution of the data. For dichotomous variables, the χ2 test was used unless any cell had an expected value of less than 5, in which case a Fisher’s exact test was employed. The Bonferroni correction was applied to adjust for potential alpha errors in the multiple comparisons. A corrected P value <0.05 was considered to be statistically significant. All statistical analyses were performed with JMP 16 software (SAS Institute, Cary, NC).

## Results

### Mapping in the Triangle of Koch and Ablation Procedure

The patient characteristics and electrophysiological features of AVNRT are shown in Table 1. The AH interval was 84±14 ms and 2 patients exhibited first-degree AV block. Of the 53 cases, an ultra-high resolution activation map in the triangle of Koch was obtained during both sinus rhythm SR and AVNRT in 27 patients (51%), during SR alone in 7 (13%), and during AVNRT alone in 19 (36%). The number of points acquired during AVNRT was 9,722 (6,214–13,212) per patient, while during SR, it was 3,355 (2,320–4,998) per patient. The earliest atrial activation (EAA) during AVNRT was observed in the anteroseptal area in 43 patients (81%) and in the midseptal area in 10 (19%). The pivot point^21,22^ was seen in 18 of the 34 patients (53%) in whom the activation map during SR was obtained.

**Table 1.** Patient and electrophysiologic characteristics. (n=53)

|  |  |
| --- | --- |
| Age (years) | 56±15 |
| Male sex | 21 (40%) |
| BMI | 23±3.9 |
| LAD (mm) | 33±5.5 |
| EF (%) | 70±6.1 |
| AH interval during sinus rhythm (ms) |  |
| Before the ablation procedure | 84±14 |
| After the ablation procedure | 86±12 |
| Electrophysiologic features of the AVNRT |  |
| TCL (ms) | 375±73 |
| AH interval (ms) | 300±83 |
| VA interval (ms) | 29±16 |
| RF ablation | 21 (40%) |

Values are the mean±SD or n (%). AH, atrio-His; BMI, body mass index; EF, ejection fraction; LAD, left atrial dimension; TCL, tachycardia cycle length; VA, ventriculo-atrial.

A fractionated potential area was observed in 19 of 34 patients (56%) during SR and in all 46 patients (100%) during AVNRT (Figure 1A-B). This area corresponded to the AVNRT termination and/or successful ablation site in all patients, with peak numbers of 8.8±1.4 during AVNRT and 5.3±1.3 during SR. Of the 32 patients who received cryoablation, cryomapping/ablation was initially applied during AVNRT in 27 patients, successfully terminating AVNRT in all (Figure 1D-E), while cryomapping/ablation was applied during SR in 5 patients. The mean number of cryomapping attempts until AVNRT termination was 2.7±2.0. Of the 27 patients, AVNRT termination was achieved during cryomapping in 22 patients and during cryoablation in 5, either due to the failure to terminate AVNRT by cryomapping (n=3) or because of the interruption of the cryomapping caused by catheter instability (n=2). The time to AVNRT termination from the initiation of the cryomapping was 12.9±3.6 seconds, and that from the initiation of the cryoablation was 13.1±4.6 seconds. However, AVNRT remained inducible despite two cryoapplications at the termination site in 5 patients. Three of those 5 patients were those in whom cryomapping did not terminate the AVNRT, but a subsequent cryoablation successfully did. In the remaining 2 patients, cryomapping terminated the AVNRT in 14.8 seconds and 19.5 seconds, respectively. The number of ablation points to achieve acute success was 3.6±1.5 in the fractionated potential-guided cryoablation (corrected P<0.05 vs. conventional RF; and P<0.05 vs. potential-guided RF), 5.4±2.1 in the fractionated potential-guided RF ablation (P=0.0825 vs. conventional RF), and 8.2±3.2 in the conventional RF ablation (Table 2).

**Table 2.** Mapping and ablation variables. (n=53)

|  | Conventional RF ablation (n=10) | Fractionated potential-guided RF ablation (n=11) | Fractionated electrogram-guided cryoablation (n=32) | P value (conventional RF vs potential-guided RF) | P value (conventional RF vs cryoablation) | P value (potential-guided RF vs cryoablation) |
| --- | --- | --- | --- | --- | --- | --- |
| **Mapping** |  |  |  |  |  |  |
| during AVNRT and SR | 2 (20%) | 6 (55%) | 19 (59%) | 0.55 | 1.00 | 1.00 |
| during SR | 2 (20%) | 1 (9%) | 4 (13%) | 1.00 | 1.00 | 1.00 |
| during AVNRT | 6 (60%) | 4 (36%) | 9 (28%) | 1.00 | 0.38 | 1.00 |
| **Ablation** |  |  |  |  |  |  |
| Junctional rhythm | 10 (100%) | 11 (100%) | 0 (0%) | NA | <0.05 | <0.05 |
| Transient AV block | 0 (0%) | 1 (9%) | 10 (31%) | 1.00 | 0.25 | 0.71 |
| 1st-degree | 0 (0%) | 1 (9%) | 5 (16%) | 1.00 | 0.94 | 1.00 |
| 2nd-degree | 0 (0%) | 0 (0%) | 3 (9%) | NA | 1.00 | 1.00 |
| 3rd-degree | 0 (0%) | 0 (0%) | 2 (6%) | NA | 1.00 | 1.00 |
| Number of cryomapping | — | — | 2.8±2.0 | NA | NA | NA |
| Number of ablations | 8.2±3.2 | 5.3±2.1 | 3.6±1.5 | 0.0825 | <0.05 | <0.05 |

Values are the mean±SD or n (%). AV, atrioventricular; AVNRT, atrioventricular nodal reentrant tachycardia; NA, not applicable; RF, radiofrequency; SR, sinus rhythm.

### Atrioventricular Block during Ablation and Dull Potentials with Continuity to the His Potential

In the anterior aspect of the triangle of Koch, dull potentials with continuity to the His potential were identified in all patients during both sinus rhythm and AVNRT, with confidence levels of 29±5% and 39±12%, respectively, and the locations of those potentials overlapped. By sliding the LAW 5 ms from the atrial activation timing on the anteroseptum to the His-bundle activation timing, those potentials were observed to propagate from the anteroseptal area to the His area in all patients (Figures 1C-D). The horizontal length of the dull potential area continuing to the His potential was 14.3±5.3 mm, and the conduction time through this area was 30±19 ms. Based on those values, the conduction velocity, calculated as the length divided by the conduction time, was 0.47±0.14 m/s.

Transient AV block occurred in 11 patients (21%): 1 during RF ablation and 10 during cryoablation. First-degree AV block occurred in 6 patients, second-degree in 3, and third-degree in 2. However, the AV block immediately resolved after the cessation of the thermal energy application in all patients. AV block was observed in 2 patients during RF ablation (n=1) and cryoablation (n=1) during SR to achieve acute success and in 6 patients during a bonus cryoapplication during SR to eliminate the AH jump and/or a single echo beat (Figure 3). In 3 other patients, AVNRT terminated with HA block (Figure 4) in 2, while AVNRT persisted despite the occurrence of AV block, suggesting a lower common pathway block (Figure 5) in 1. The ablation site causing AV block corresponded to the EAA site during AVNRT in 6 patients. All AV block sites overlapped with the dull potential area in the phase just before the His potential during AVNRT and/or SR, with a confidence setting of ≥24% (35 [24-60]%). Conversely, in 42 patients without AV block, ablation was not performed in this area. Given that, the ablation in the dull potential area highlighted by ≥24% during SR and AVNRT had a potential risk of developing AV block with a sensitivity of 100% and specificity of 100%. The successful ablation site was immediately below the highlighted area, where no dull potential was recorded in any patients. The successful ablation site was located 7.0 ± 3.3 mm below the dull potential area during AVNRT and 6.2 ± 2.6 mm away during SR.

**Figure 3.**
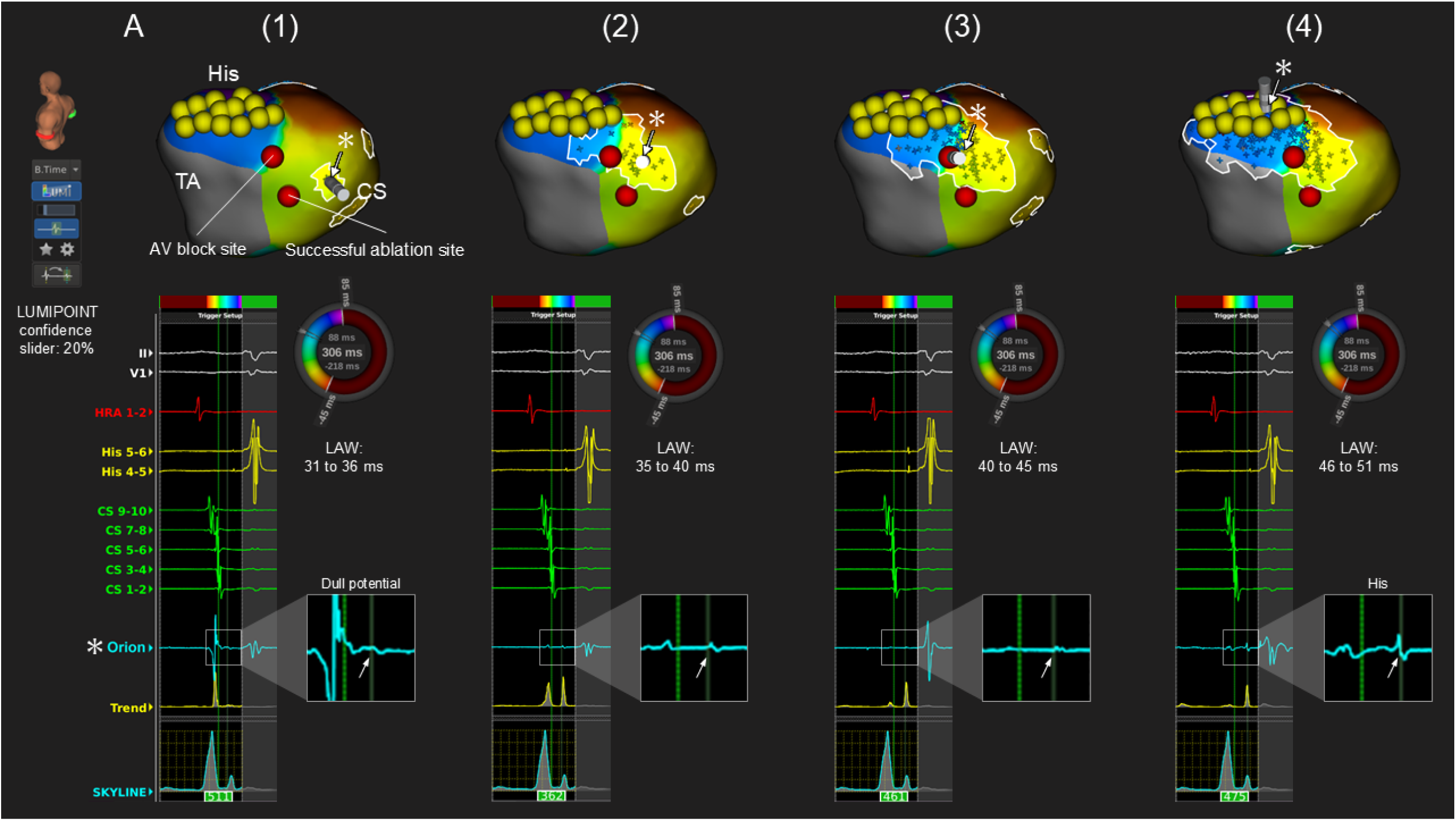
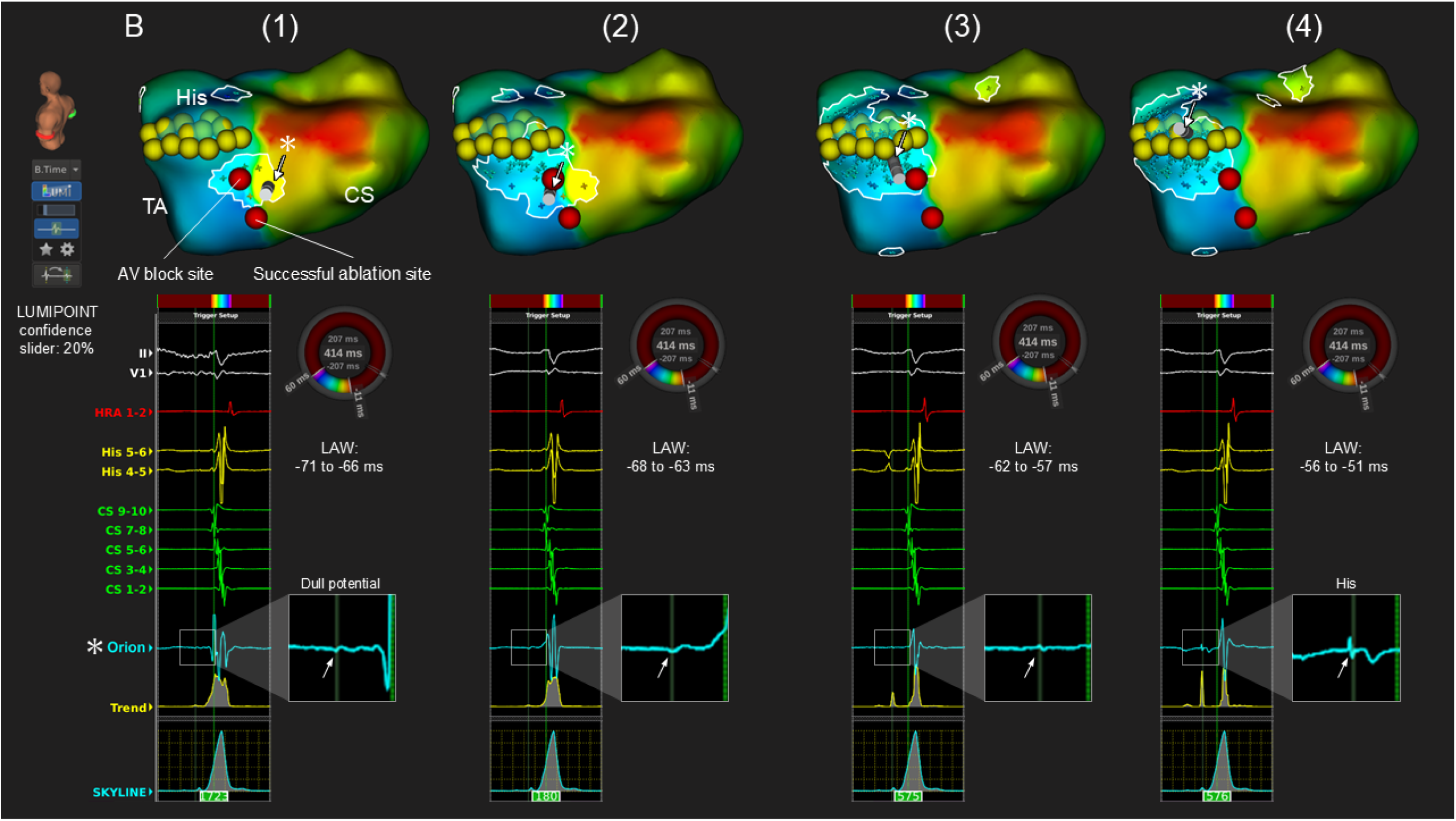
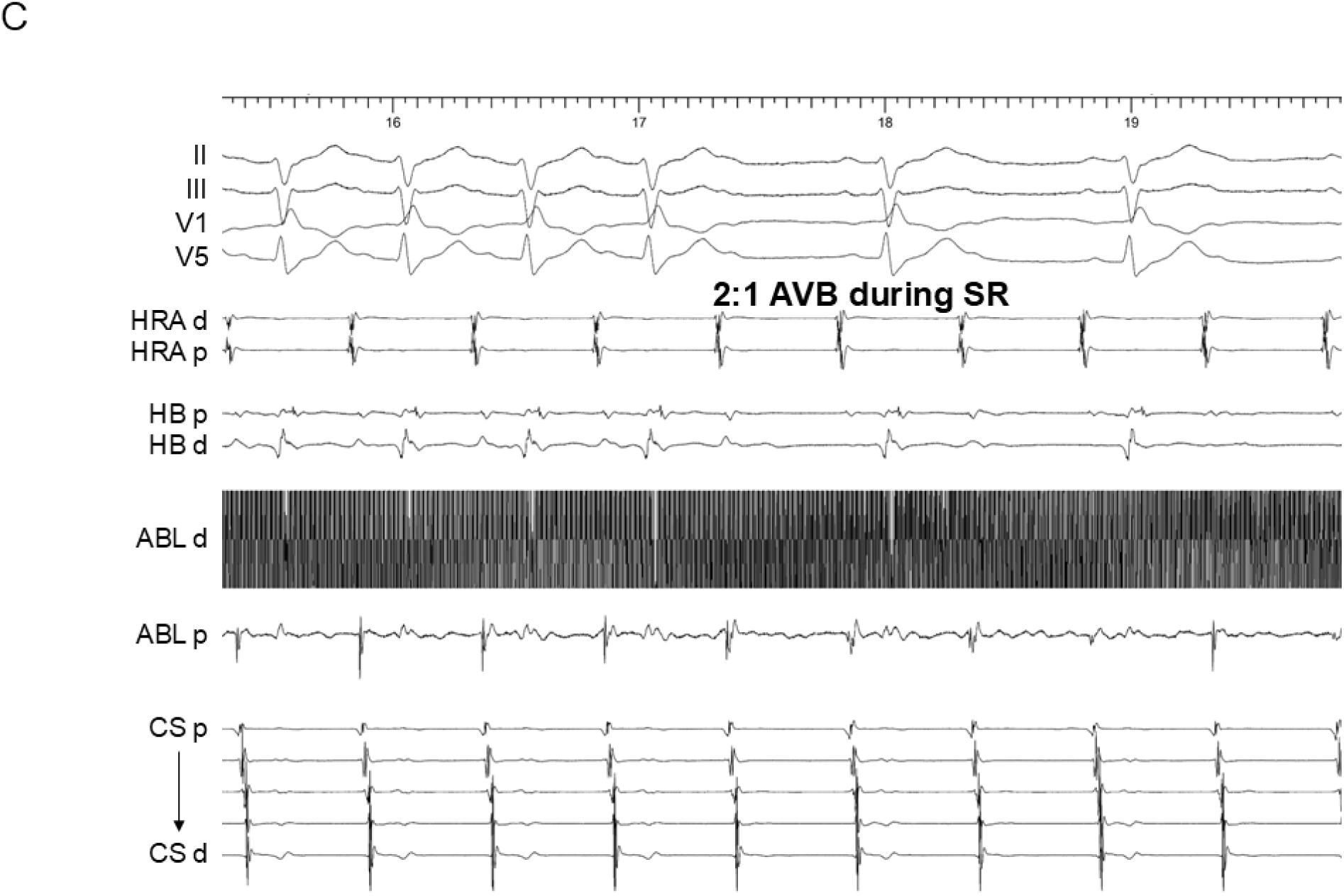
An activation map and local electrograms in the triangle of Koch in a patient where the cryoablation during SR caused 2:1 atrioventricular block. **(A)** The propagation of the annotated dull potentials during SR with confidence levels of 20% and **(B)** during AVNRT with confidence levels of 20%. By sliding the local activation window (LAW) by 5 ms, those potentials are observed propagating from the anteroseptal area to the His region. **(C)** Intracardiac electrograms of 2:1 atrioventricular block during cryoablation. The abbreviations are as in Figure 1.

**Figure 4.**
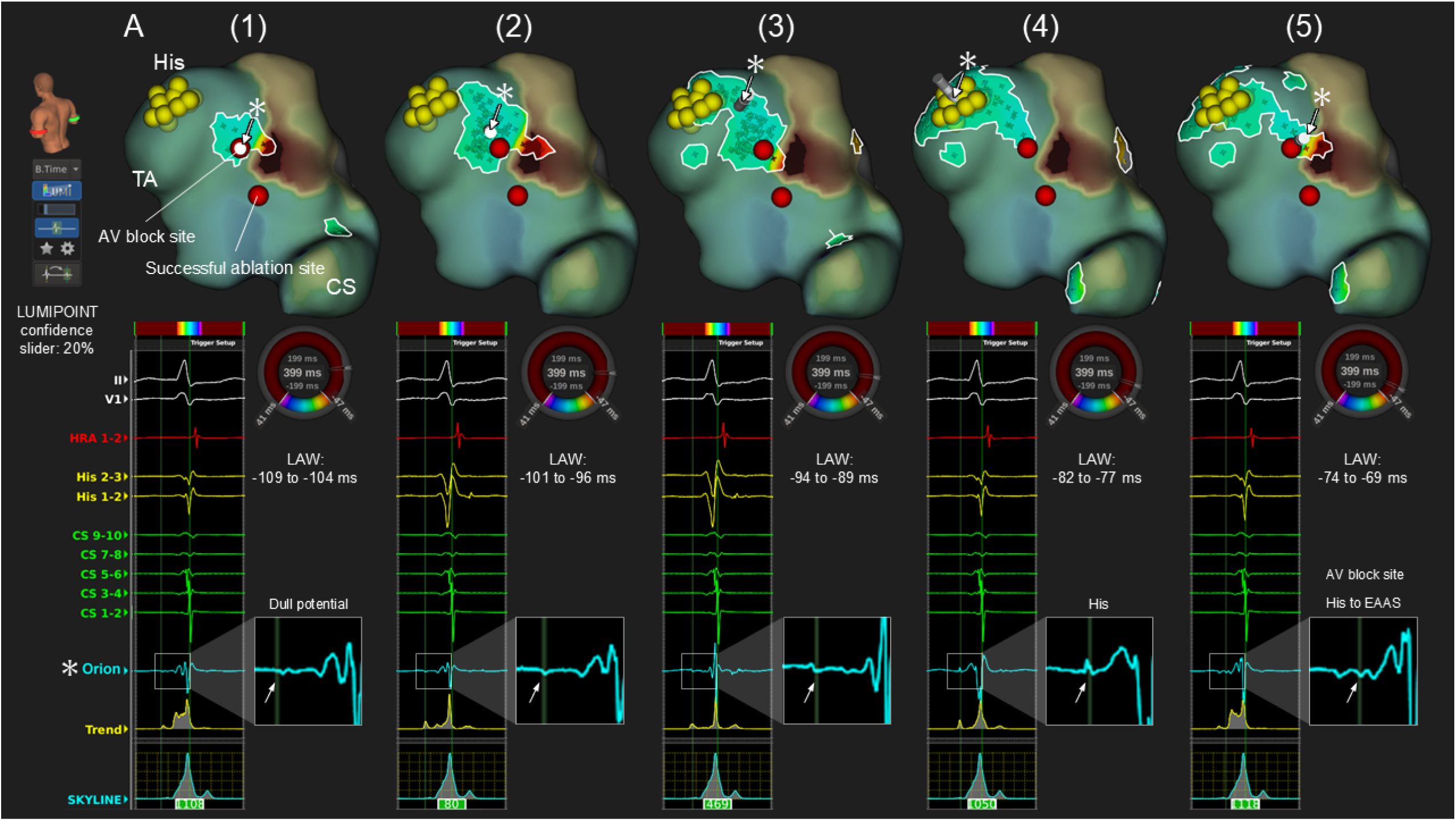
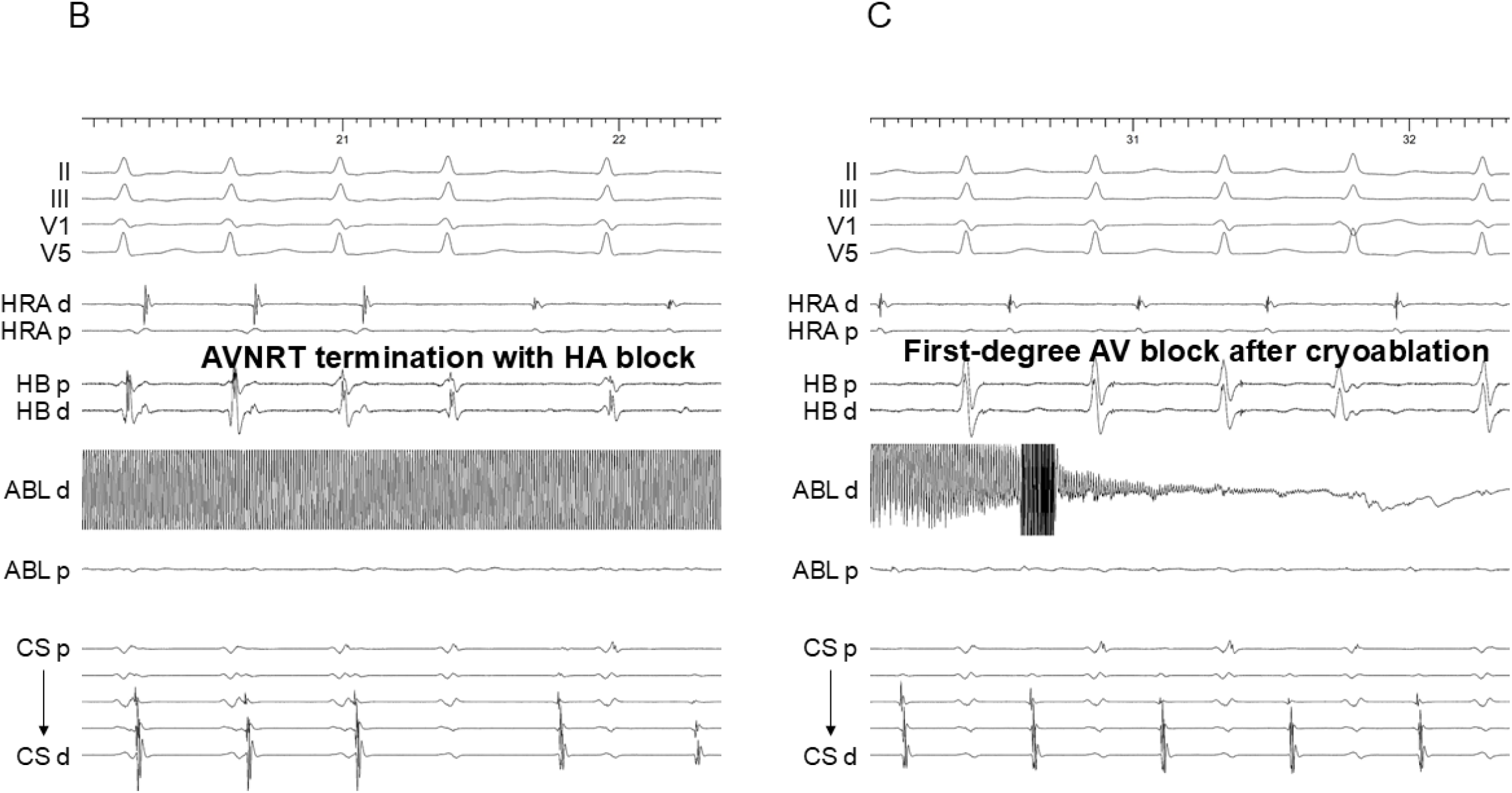
An activation map and local electrograms in the triangle of Koch in a patient where the cryoablation terminated AVNRT with His-atrial block and caused first-degree atrioventricular block. **(A**) The propagation of the annotated dull potentials during SR with confidence levels of 20%. By sliding the local activation window (LAW) by 5 ms, those potentials are observed propagating from the anteroseptal area to the His region. **(B)** Intracardiac electrograms of AVNRT termination with His-atrial block and **(C)** first-degree atrioventricular block. The abbreviations are as in Figure 1.

**Figure 5.**
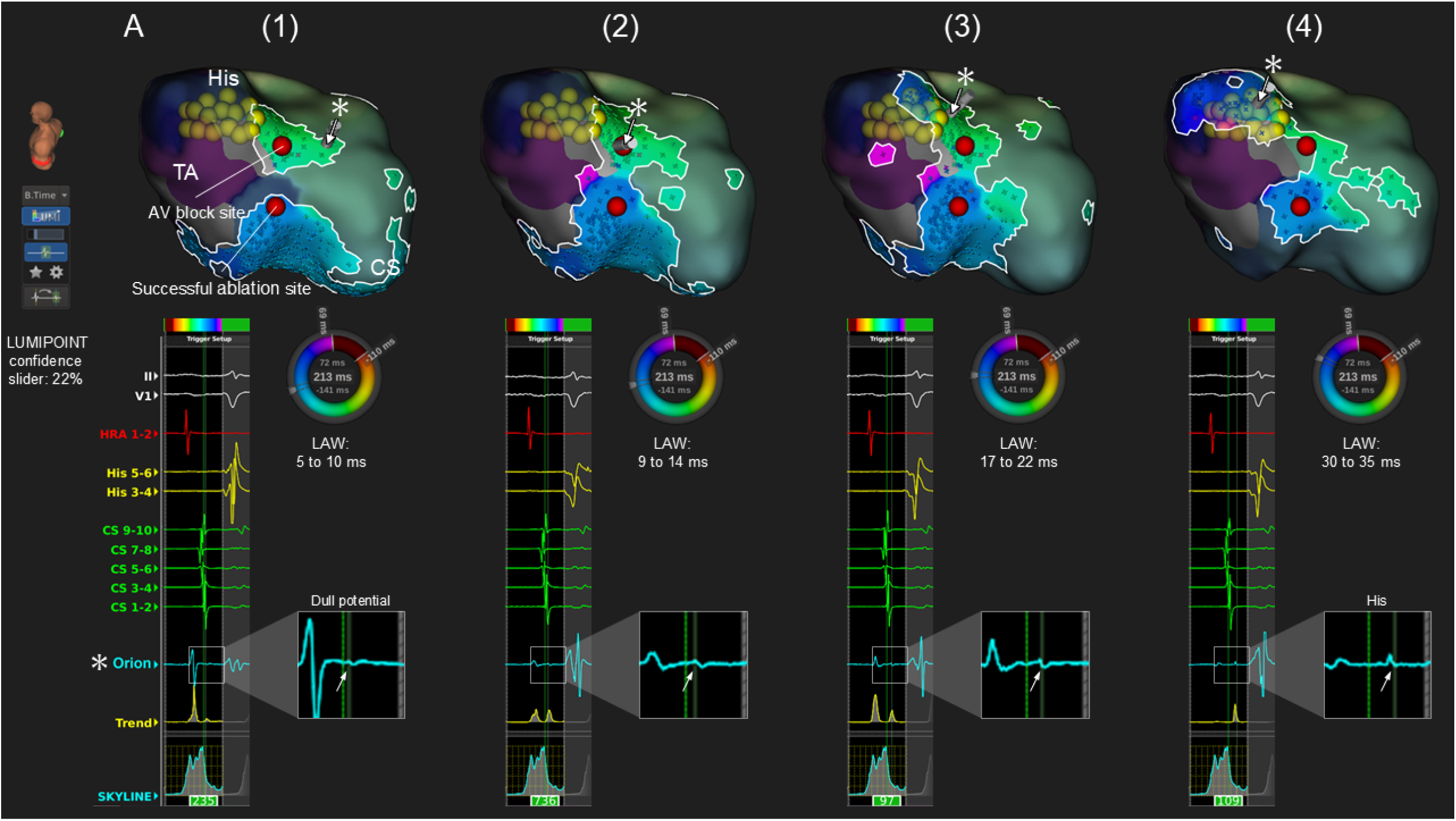
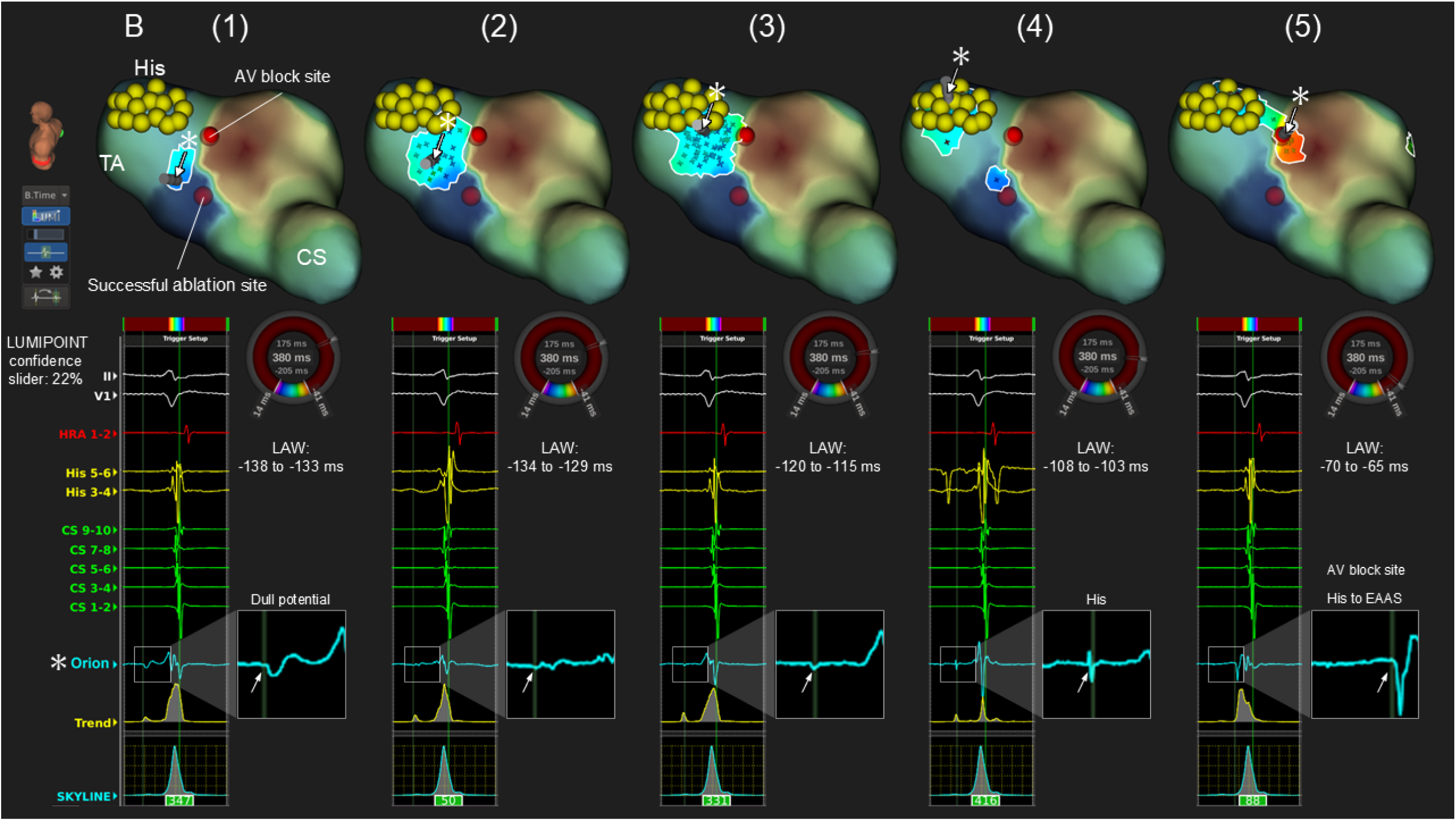
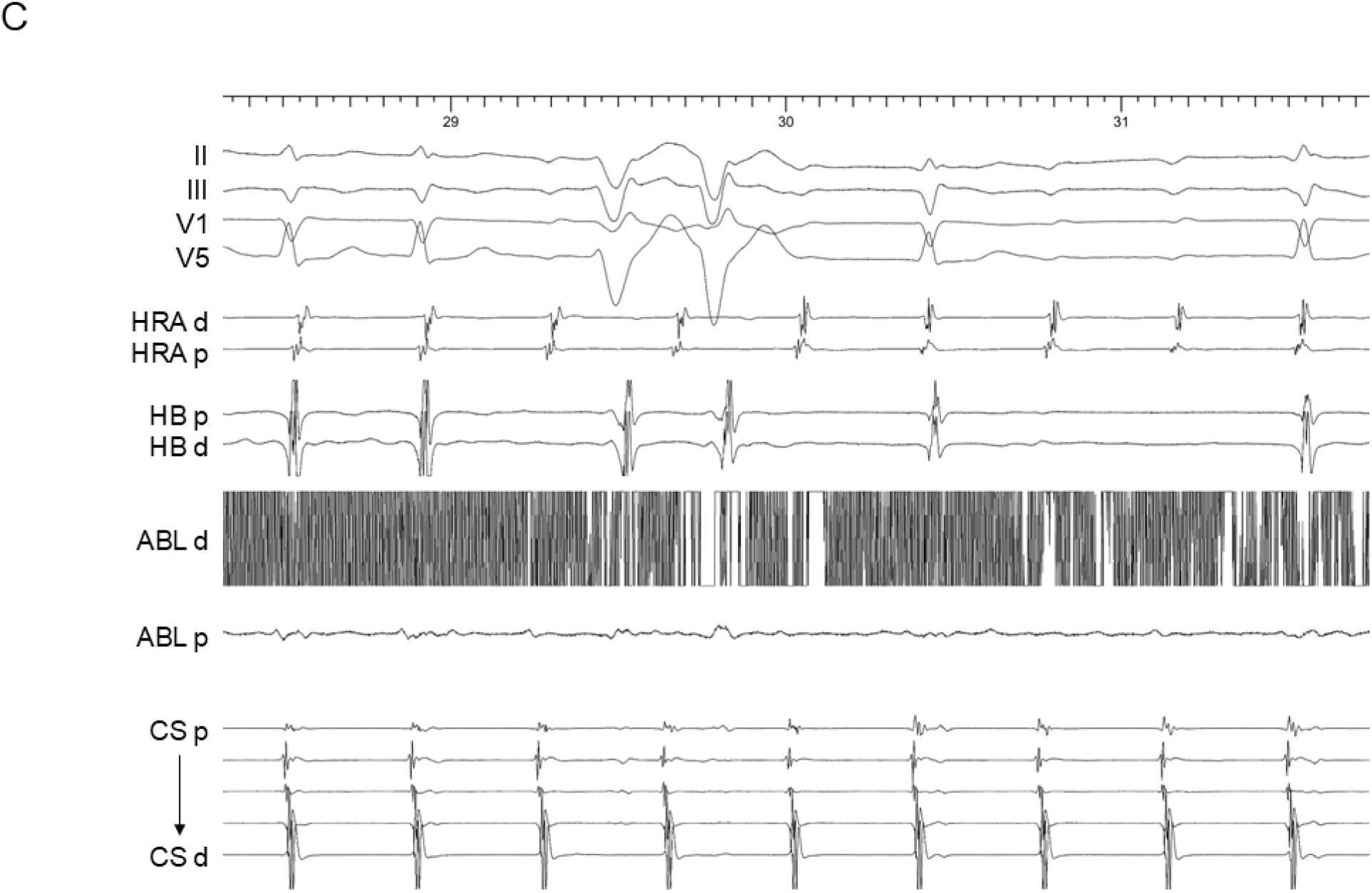
An activation map and local electrograms in the triangle of Koch in a patient where AVNRT persisted despite the occurrence of lower common pathway block during cryoablation. **(A)** The propagation of the annotated dull potentials during SR with confidence levels of 22% and **(B)** during AVNRT with confidence levels of 22%. By sliding the local activation window (LAW) by 5 ms, those potentials were observed propagating from the anteroseptal area to the His region. **(C)** Intracardiac electrograms of sustained AVNRT despite the occurrence of atrioventricular block. The abbreviations are as in Figure 1.

### Ablation Outcomes and Follow-up

Acute success was achieved in all patients: complete SP elimination in 27 patients (51%), SP modulation with residual dual AVN physiology without any echo beats in 9 patients (17%), and with a single echo beat in 17 patients (32%). During a median follow-up of 6 months (6-12 months), an AVNRT recurrence occurred in 1 patient who received a fractionated potential-guided RF application strategy.

## Discussion

The main findings of this study were as follows: (1) the number of ablation points in the fractionated potential-guided cryoablation strategy targeting AVNRT termination was fewer than that with RF ablation, and (2) the RF/cryo application in the area where dull potentials with continuity to the His potentials during SR and AVNRT were detected, with a confidence setting of ≥24% [35 (24-60)%], posed a risk of AV block.

### Fractionated potential-guided cryoablation strategy terminating AVNRT

Our previous study revealed that the SP area was highlighted using the LUMIPOINT module in 83% of AVNRT cases by acquiring potentials during AVNRT, and those areas corresponded to 90% of the sites of successful ablation.^3^ The highlighted area was located near the entrance of the SP, as evidenced by the resetting of AVNRT by an atrial extrastimulus at the site with the lowest pacing prematurity. Those findings suggested that the fractionated potential may reflect the participation of tissue at the insertion of the SP into the atrium, specifically the transitional cells. Furthermore, the SP potential-guided ablation strategy for slow-fast AVNRT may help prevent redundant RF applications without reducing the success rate.

In the next phase of our research, we evaluated SP potential-guided cryoablation targeting the termination of AVNRT. The number of ablation points in this strategy was fewer than that in the fractionated potential-guided RF ablation and conventional anatomical RF ablation. Even in previous studies on cryoablation, the number of ablation points appeared to be lower with the AVNRT termination strategy (a median of 4.2 cryoapplications) than conventional anatomical approach (a median of 4.5-7 cryoapplications).^7–9,11^ Considering our result of 3.6±1.5 cryoapplications, the SP potential-guided cryoablation targeting AVNRT termination may further reduce the number of ablation points. Furthermore, the number of cryomapping sites appears to have been higher in the previous studies (4.3-7 sites) than in our study with 2.8±2.0 sites. This may be attributed to the higher specificity of the ablation target with this approach. From this perspective, our strategy could offer an advantage of reducing both the cryomapping and cryoablation points.

Additionally, the time to AVNRT termination was a strong predictor of a successful cryoablation. In this study, the time to successful AVNRT termination without an acute recurrence was ≤14.8 seconds in all patients, suggesting that an AVNRT termination time of ≤15 seconds might predict the absence of acute recurrence after cryoablation. This value is also consistent with the findings from previous research, which reported a termination time of 20 seconds.^11^

### Dull Potentials with Continuity to the His Potential

During both SR and AVNRT, tiny, dull potentials with continuity to the His potentials were identified in the anteroseptal area, with a confidence level of ≥24% (35[24-60] %). Additionally, in all 11 patients who experienced transient AV block, the block occurred during RF or cryoablation applications in this region. Conversely, in 42 patients without AV block, ablation was not performed in this area. Those observations suggest that those dull potentials may represent the cAVN potential extending to the proximal His-bundle potential.

The cAVN primarily consists of slender, interwoven transitional cells with poor cell-to-cell coupling, resulting in its slow conduction properties. In contrast, the proximal His bundle consists of interweaving Purkinje-type cells, and as the bundle progresses, it becomes predominantly composed of large, longitudinally oriented Purkinje-type cells. This difference in the cellular composition explains the slow conduction over the cAVN of 0.12 m/s,^23^ and rapid conduction over the His bundle of 1.5 m/s.^24^ This disparity explains the low-amplitude, low-frequency hump-shaped atrial potential of the cAVN, and the high-frequency potential of the His bundle, which are consistent with the characteristics of the dull potentials observed in our study.^5^ The conduction velocity of the dull potential area in our study was calculated as 0.47±0.14 m/s, which was approximately intermediate between the conduction velocities of the cAVN and His bundle reported in previous studies. In 2 patients, cryoablation in this area terminated the AVNRT with HA block, which indicated that those dull potentials likely represented the cAVN rather than the proximal His-bundle. However, the dull potentials still failed to encompass the entire phase of AVNRT, particularly during the diastolic period. From that perspective, the cAVN may not be fully delineated.

The propagation of the dull potentials was easily discernible during SR but not during AVNRT, possibly due to the difference in the activation sequence between the two rhythms. During SR, the activation proceeded sequentially through the atrial myocardium, FP, cAVN, and His-bundle, whereas during AVNRT, the activation from the cAVN to the His-bundle and from the retrograde FP conduction to the atrial myocardium occurs simultaneously. However, during SR, the dull potentials overlapped with the surrounding atrial activation. In contrast, during AVNRT, the timing of the dull potentials was distinct from the atrial activation. Therefore, the propagation during SR may be more useful for understanding cAVN and His-bundle conduction, while that during AVNRT may be more advantageous for accurately identifying the location of the cAVN. Nonetheless, in one case, a lower common pathway block occurred during a cryoapplication in the dull potential area, suggesting that differentiating the lower common pathway from the cAVN may be challenging, as these structures could be histologically unique.

### The Proposed Ablation Strategy Potentially Reducing Redundant Applications and Complications

Based on our results, we recommend performing activation mapping during AVNRT. The first step with this activation map is the visualization of the SP area with fractionated potentials, which are highlighted by the LUMIPOINT complex activation module. The number of peaks was initially set to 8.4 and was then adjusted to ensure a highlighted area of approximately 1.0 cm² for each patient. The next step was the visualization of the cAVN-His area, highlighting the dull potentials with continuity to the His potentials, using a confidence setting of 24-60%. The ablation target should be determined as the fractionated potential area located 7-10 mm below the inferior edge of the cAVN-His potential area where the dull potentials were not recorded. Cryomapping was initiated in this area during AVNRT. If the AVNRT was terminated within 15 seconds, cryoablation should be performed using freeze-thaw-freeze applications.

## Conclusion

The fractionated potential-guided cryoablation strategy targeting AVNRT termination required fewer cryoapplications than RF ablation. The RF/cryo applications in the area where dull potentials with continuity to the His potentials during SR and AVNRT were detected, with a confidence setting of ≥24% [35 (24-60)%], posed a risk of AV block.

## Data Availability

The datasets generated and analyzed during the current study are available from the corresponding author on reasonable request.

## CONFLICT OF INTEREST

Authors declare no conflict of interest for this article.

## ETHICS STATEMENT

Approval of research protocol: the study was approved by the Institutional Review Board of Nihon University Itabashi Hospital. Approval number: RK-241008-2.

## PATIENT CONSENT STATEMENT

An opt-out system was used to obtain the patients’ content for the use of their clinical data for research purposes.

## CLINICAL TRIAL REGISTRATION

N/A.

## ACKNOWLEDGEMENTS

We would like to extend our heartfelt thanks to Mr. John Martin for his assistance with the English language editing. I would also like to express my deepest gratitude to my co-authors and the patients who contributed to this work.

## Notes

### Competing Interest Statement

The authors have declared no competing interest.

### Author Declarations

The study was approved by the Institutional Review Board of Nihon University Itabashi Hospital. Approval number: RK-241008-2.

